# Serum anti-Spike antibody titers before and after heterologous booster with mRNA-1273 SARS-CoV-2 vaccine following two doses of inactivated whole-virus CoronaVac vaccine

**DOI:** 10.1101/2021.12.24.21268360

**Authors:** Robert Sinto, Dwi Utomo, Suwarti, Erni J. Nelwan, Henry Surendra, Cindy Natasha, Fransiska, Deborah Theresia, Adella Faiqa Ranitria, Decy Subekti, Nunung Nuraeni, Winahyu Handayani, Mutia Rahardjani, J. Kevin Baird, Susanna Dunachie, Anuraj H. Shankar, Raph L. Hamers

**Affiliations:** Faculty of Medicine, Universitas Indonesia, Jakarta, Indonesia; Cipto Mangukusumo National Hospital, Jakarta, Indonesia; St Carolus Hospital, Jakarta, Indonesia; Centre for Tropical Medicine and Global Health, Nuffield Department of Medicine, University of Oxford, Oxford, UK; Pasar Minggu Hospital, Jakarta, Indonesia; Eijkman-Oxford Clinical Research Unit, Jakarta, Indonesia; Infectious Disease and Immunology Research Cluster, Indonesian Medical Education and Research Institute, Jakarta, Indonesia; Mahidol-Oxford Tropical Medicine Research Unit, Mahidol University, Bangkok, Thailand

**Keywords:** SARS-CoV-2, COVD-19, inactivated vaccine, CoronaVac, mRNA-1273, antibodies

## Abstract

**Background:** The inactivated whole-virus vaccine CoronaVac (SinoVac) is the COVID-19 vaccine most administered worldwide. However, data on its immunogenicity and reactogenicity to heterologous boosting with mRNA vaccines are lacking.

**Methods:** In a cohort of hospital staff in Jakarta, Indonesia, who received two-dose CoronaVac six months prior (median 190 days, IQR165-232), we measured anti-Spike IgG titers on paired serum samples taken before and 28 days after a 100μg mRNA-1273 (Moderna) booster. We performed correlations and multivariable ordinal regressions.

**Findings:** Among 304 participants, the median age was 31 years (range 21-59), 235 (77.3%) were women, 197 (64.8%) had one or more previous SARS-CoV-2 infections (including 155 [51.0%] who had a post-CoronaVac breakthrough infection. Pre-boost IgG titers correlated negatively with the time since the latest documented “virus exposure” (either by the second CoronaVac or SARS-CoV-2-infection whichever most recent). Previous SARS-CoV-2 infection and a longer time interval between second vaccine and mRNA-1273 boost were associated with a higher pre-boost IgG titer. Post-booster, the median IgG titer increased 9.3-fold, from 250 (IQR32-1389) to 2313 (IQR1226-4324) binding antibody units (BAU/mL) (p<0.001). All participants, including seven whose pre-boost IgG was below assay detection limits, became seropositive and all reached a substantial post-boost titer (≥364 BAU/mL). Post-boost IgG was not associated with pre-boost titer or previous SARS-CoV-2 infection. Booster reactogenicity was acceptable, with 7.9% of participants experiencing short-lived impairment of activities of daily living (ADL).

**Interpretation:** A heterologous, high-dose mRNA-1273 booster after two-dose CoronaVac was highly immunogenic and safe, including in those most in need of improved immunity.

**Funding:** Wellcome Trust, UK

**Research in context:** *Evidence before this study:* The inactivated whole-virus vaccine CoronaVac (SinoVac) is the COVID-19 vaccine most administered worldwide, at around 2 billion doses in 54 countries. Concerns that CoronaVac has lower immunogenicity than virus vector or mRNA vaccines, with pronounced decreases of neutralising antibody titres within a few months, and reduced effectiveness in the older population, highlight the urgent need for immunogenic, safe and well-tolerated booster schedules, especially with Omicron rapidly emerging. We used the terms “SARS-CoV-2”, “COVID-19”, “vaccine”, “booster” to search PubMed and medRxiv up to Dec 22th, 2021, with no language or date restrictions, to identify clinical trials and real-world studies reporting on the immune responses and reactogenicity to a “third booster” of currently approved COVID-19 vaccines. Previous research reported that neutralising antibody responses elicited by all currently approved vaccines (mRNA, adenovirus-vectored, inactivated, and protein subunit) declined to varying degrees after 6-8 months after full-schedule vaccination. Several clinical trials have evaluated heterologous (“mix and match”) vaccination schedules, demonstrating robust immune responses in adults. After two-dose CoronaVac, BNT162b2 (Pfizer-BioNTech) boost was significantly more immunogenic than a homologous booster against wild-type and Variants of Concern (VOCs) Beta, Gamma and Delta, and AZD1222 boost increased spike RBD-specific IgG 9-10-fold, with high neutralizing activity against the wild type and VOCs. Compared to previous SARS-CoV-2 variants, current vaccine boosters appeared to neutralise Delta to a slightly lesser degree, and Omicron to a substantially lesser degree, although preliminary data from Moderna found that the authorised dose (50μg) of the mRNA-1273 boost increased antibodies 37-fold and the high-dose (100μg) boost 83-fold.

*Added value of this study:* To our knowledge, this study is the first to provide critical real-world evidence that heterologous boosting with high-dose mRNA-1273 vaccine after CoronaVac is highly immunogenic, safe and well-tolerated in adults. After a primary course of two-dose CoronaVac, we found that a high-dose (100μg) mRNA-1273 booster was immunogenic for all participants in a highly exposed cohort of hospital staff in Jakarta, Indonesia, in the context of Delta predominance, particularly for those with the lowest pre-boost antibody levels. All participants became seropositive and all reached a substantial post-boost titer (≥364 BAU/mL), up to a median 9.3-fold increase. Booster reactogenicity was acceptable, with 7.9% of participants experiencing short-lived impairment of activities of daily living

*Implications of all the available evidence:* The study findings contribute to informing policy makers on flexible options in deploying COVID-19 vaccines in mix-and-match schedules, with particular relevance for countries that are largely dependent on inactivated vaccines. Further trials are warranted that assess clinical endpoints of optimized doses of mRNA-1273 booster, and variant-specific or multivalent vaccines in response to decreased protection against emerging SARS-CoV-2 VOCs.

## Introduction

Vaccine-induced population immunity is a key global strategy to control the COVID-19 pandemic, and to date eight COVID-19 vaccines have received Emergency Use Listing (EUL) by the WHO^1^. Although most studies suggest well preserved protection against severe COVID-19 disease and death, including against variants such as the Delta (B.1.617.2) strain, accumulating evidence shows a progressive decline in protection after a two-dose vaccine schedule in preventing breakthrough infections associated with diminishing humoral immunity over time^2–5^. In addition, neutralisation and vaccine effectiveness after two-dose vaccine schedules are greatly reduced for the recently emerged Omicron variant (B.1.1.529)^6^, with significant restoration after a third “booster dose” of an mRNA vaccine^7^. Recent trials suggested that heterologous (or “mix and match”) virus-vectored or mRNA booster strategies were more immunogenic than a homologous schedule^8–12^, albeit with increased reactogenicity in some combinations^13^.

CoronaVac (SinoVac Life Sciences Co Ltd, Beijing, China), an inactivated whole-virus vaccine, received WHO EUL on June 1^st^, 2021^14^, and is currently the COVID-19 vaccine with most doses administered worldwide at around 2 billion in 54 countries, as per December 20^st^, 2021. Vaccine efficacy against symptomatic illness of two-dose CoronaVac was reported between 50.5% and 83.5% in trials^14,15^, and 65.9% and 49.4% in real-world studies in Chile^16^and Manaus, Brazil, respectively^17^. Real-world data from Chile reported high prevention of hospitalisation (87.5%), ICU admission (90.3%), and COVID-19-related death (86.3%)^16^. However, recent studies reported CoronaVac to have lower immunogenicity than virus vector or mRNA vaccines^18,19^, and a pronounced decrease of neutralising antibody titres within a few months^19–21^, and reduced effectiveness in the elderly ^22,23^. This lack of protection is concerning, especially given the rapid emergence of Omicron^6^.

Indonesia, with the world’s fourth largest population (270 million) has the second highest number of COVID-19 cases (6.26 million) and deaths (144.000) in Asia (per December 20^th^, 2021)^24^. The countrywide mass vaccination campaign, launched in January 2021, has been predominantly based on two-dose CoronaVac. The increased transmission of Delta variant during Indonesia’s second COVID-19 surge in July 2021^25^, including among dual-vaccinated health care workers, prompted the Ministry of Health in August 2021 to offer a heterologous, high-dose (100μg) booster of the mRNA-1273 vaccine (Moderna Inc., Cambridge, US) to frontline health care workers.

Given the dearth of data on effectiveness, immunity and reactogenicity of a heterologous booster dose for inactivated whole-virus COVID-19 vaccines^18,26^, we investigated the immunogenicity and reactogenicity of a high-dose (100μg) heterologous mRNA-1273 booster given six months after two-dose CoronaVac to heavily SARS-CoV-2 exposed hospital staff, measuring anti-spike (anti-S) IgG antibody titers before and one month after the booster, and the occurrence and severity of adverse events within seven days.

## Methods

### Design and population

The Indonesia Vaccine Immunity & Infection Evaluation (INVITE) study is a longitudinal observational cohort of hospital staff aged 18 years or older in St Carolus and Pasar Minggu hospitals in Jakarta who received the 100μg mRNA-1273 booster dose. The present analysis includes all study participants for whom a paired pre-and post-booster serum sample was available. The study was approved by the research ethics committee of Faculty of Medicine Universitas Indonesia (841/UN2.F1.ETIK/PPM.00.02/2021) and Oxford Tropical Research Ethics Committee (22-21). All participants provided written informed consent.

### Procedures

Basic demographic and clinical data were captured on an online case report form. Solicited signs of local or systemic reactogenicity during the seven days following the booster dose were recorded in a daily patient diary, and, after study physician verification, were graded for severity (mild, moderate, severe, or disrupting activities of daily living [ADL]). Past COVID-19 was defined as a previously documented PCR-confirmed SARS-CoV-2 infection. Venous blood samples were drawn on the day of the booster dose (between August 5^th^, and October 15^th^, 2021) and 28 days (allowed window +10 days) thereafter (between September 13^th^ and November 12^th^, 2021).

Serum was stored at -80°C and transferred to a commercial laboratory, where titers of IgG antibodies against the SARS-CoV-2 spike receptor-binding domain were determined using the chemiluminescent microparticle immunoassay (CMIA) SARS-CoV-2 IgG II Quant assay (Abbott Laboratories, Abbott Park, IL, US) on the Architect i2000sr platform, in accordance with the manufacturer’s package insert. Samples with results above the upper limit of quantification were tested again after dilution. The strength of the response (in relative light units) was determined relative to an IgG concurrent calibrator, and reflects the quantity of IgG antibodies present. We expressed test results in WHO International Standard binding antibody units (BAU)/mL using the manufacturer’s conversion factor (1 BAU/mL = 0.142 x arbitray units[AU]/mL)^27^. Seropositivity was defined as ≥7.1 BAU/mL.

### Statistical analysis

Paired comparisons of IgG titers before and after mRNA-1273 booster dose were performed using the Wilcoxon matched pairs signed-rank test. Correlations of log_10_ IgG titers were expressed using Spearman coefficient (r_s_). Multivariable ordinal (proportional odds) logistic regression was performed to assess factors associated with pre-and post-boost log_10_-transformed IgG titer (caterogised in quartiles). Multivariable logistic regression was performed to assess factors associated with occurrence of any severe or disrupting adverse reactions. The independent variables included in the analysis were age, sex, previous SARS-CoV-2 infection, timing of previous SARS-CoV-2 infection (before or after two-dose CoronaVac), presence of any comorbidity (i.e. cardiovascular disease including hypertension, obesity [BMI>30 kg/m^2^], diabetes mellitus, asthma), pre-boost IgG titer, time interval between first and second CoronaVac dose, time interval between second CoronaVac and mRNA-1273 booster dose. A two-sided *P* <0.05 was considered significant. Statistical analysis was performed with Stata/IC 15.1 (StataCorp, College Station, TX, USA).

## Results

### Participants’ characteristics

There were 304 (86.1%) of 353 cohort participants with complete pre-and post-booster sample pairs, who were included in the analysis. The median age was 31 years (IQR, 27-44, range 21-59), 235 (77.3%) were women, and 28 (9.2%) had one or more comorbidities, including obesity, cardiovascular disease, diabetes mellitus, and asthma (**Table 1**). There were 197 (64.8%) with previous SARS-CoV-2-infection, including 21 (6.9%) with two infections, and 155 (51.0%) breakthrough infections post-CoronaVac. The mRNA-1273 booster was given at a median of 190 days (IQR 165-232) after the second CoronaVac dose. After the mRNA-1273 booster, the overall median IgG titer showed a 9.3 fold increase, from 250 (IQR 32-1389) to 2313 (IQR 1226-4324) BAU/mL (p< 0.001), and seropositivity increased from 96.4% (293) to 100% (304). (**Table 1**). **Table S1** and **Figures S2-3** summarize the pre-post change in median IgG titers for participant subgroups.

**Table 1.** Participants’ characteristics (n=304)

| Characteristic | Total<br>(N=304) | Naïve<br>(N=107) | Previously SARS-CoV-2 infected<br>(N=197) |
| --- | --- | --- | --- |
| Age, median (IQR) - yrs | 31 (27-44) | 30 (26-43) | 31 (27-45) |
| 21-29 | 134 (44.08) | 52 (48.60) | 82 (41.62) |
| 30-39 | 68 (22.37) | 21 (19.63) | 47 (23.86) |
| 40-49 | 57 (18.75) | 23 (21.50) | 34 (17.26) |
| 50-59 | 45 (14.80) | 11 (10.28) | 34 (17.26) |
| Sex |  |  |  |
| Women | 235 (77.30) | 83 (77.57) | 152 (77.16) |
| Men | 69 (22.70) | 24 (22.43) | 45 (22.84) |
| Previous SARS-CoV-2 infection episode |  |  |  |
| 0 | 107 (35.20) | - | - |
| 1 | 176 (57.89) | - | - |
| 2 | 21 (6.91) | - | - |
| Previous SARS-CoV-2 infection timing |  |  |  |
| None | 107 (35.20) | - | - |
| Before CoronaVac vaccination | 42 (13.82) | - | - |
| After CoronaVac vaccination | 155 (50.99) | - | - |
| Comorbidities |  |  |  |
| Yes | 61 (20.07) | 19 (17.76) | 42 (21.32) |
| No | 243 (79.93) | 88 (82.24) | 155 (78.68) |
| Obesity | 39 (12.87) | 13 (12.15) | 26 (13.27) |
| Cardiovascular disease (including hypertension) | 20 (6.58) | 6 (5.61) | 14 (7.11) |
| Asthma | 7 (2.30) | 2 (1.87) | 5 (2.54) |
| Diabetes mellitus | 4 (1.32) | 2 (1.02) | 2 (1.87) |
| Data before mRNA-1273 booster dose (Day 0) |  |  |  |
| Interval between first and second dose, days (median, IQR) | 14 (14-17) | 14 (14-17) | 14 (14-18) |
| Interval between second dose and booster, days (median, IQR) | 190 (165-232) | 177 (151-190) | 213 (171-239) |
| IgG titer, median (IQR) - BAU/mL | 250 (32-1389) | 32 (15-84) | 736 (146-1868) |
| Seropositivity % | 293 (96.38) | 97 (90.65) | 196 (99.49) |
| Data after mRNA-1273 booster dose (Day 28) |  |  |  |
| Interval between booster and Day 28 IgG measurement, median (IQR) | 34 (31-36) | 35 (34-39) | 31 (30-35) |
| IgG titer Day 28, median (IQR) - BAU/mL | 2313 (1226-4324) | 2834 (1228-4687) | 2132 (1201-3855) |
| Seropositivity % | 304 (100) | 107 (100) | 197 (100) |

Abbreviations: BAU, binding antibody units (according to WHO International Standard) Seropositivity was defined as $\geq 7.1$ binding antibody units (BAU)/mL (WHO International Standard)

### Correlations of pre and post mRNA-1273 booster IgG titers

Pre-boost IgG titers varied from 0.97 BAU/mL to 21 466 BAU/mL (4.35 log_10_ range), and were higher overall in previously infected than in naïve participants (**Figure 1** and **Figure S1**). Pre-boost IgG titers correlated negatively with the time since the most recent documented “virus exposure”, either by the second CoronaVac or SARS-CoV-2 infection (whichever most recent). Participants with a breakthrough infection after post-Coronavac had the shortest time since most recent “virus exposure”, and had a higher pre-boost IgG titer than those infected before CoronaVac and naïve participants. Participants with a breakthrough infection post-CoronaVac had the shortest time since most recent “virus exposure” and a higher pre-boost IgG titer than the naïve participants.

**Figure 1.**
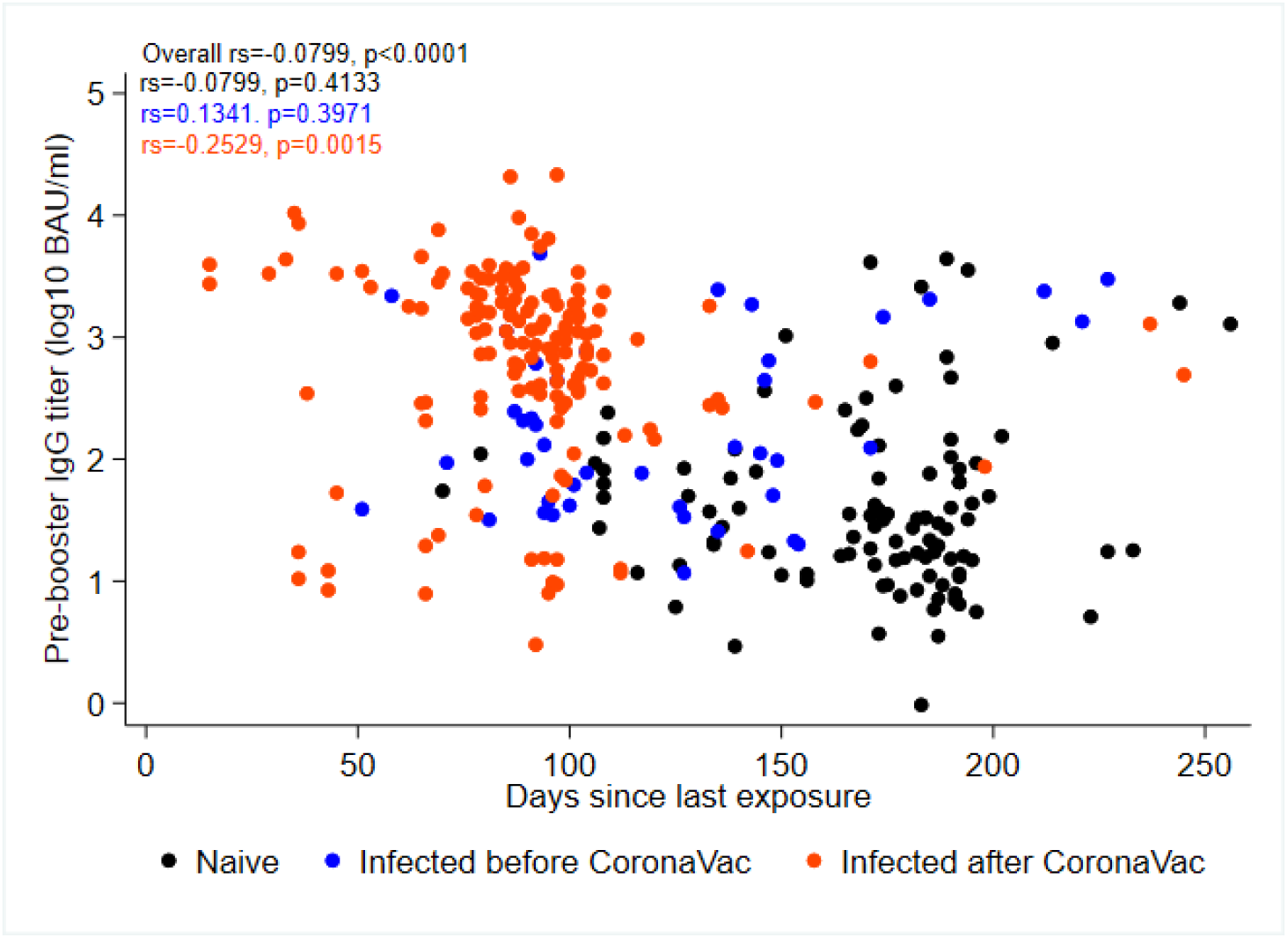
Scatter plot of anti-Spike IgG titers before mRNA-1273 booster and time since most recent “virus exposure”. Pre-boost titers were higher overall in previously infected than in naïve participants. Pre-boost IgG titers correlated negatively with the time since the most recent documented “virus exposure” (either by the second CoronaVac dose or SARS-CoV-2 infection whichever most recent). Participants with a breakthrough infection post-Coronavac had the shortest time since most recent “virus exposure”, and had a higher pre-boost IgG titer than those infected before Coronavac and naïve participants.

All participants reached a substantial post-boost titer, ranging between 364 and 195 452 BAU/mL (2.73 log_10_ range), and this was irrespective of the pre-boost IgG titer, previous infection, or the timing of the previous infection (either before or after two-dose CoronaVac vaccination). Participants with a breakthrough infection after two-dose CoronaVac had a higher pre-boost IgG titer than those infected before Coronavac and the naïve participants. Post-boost IgG titers were in the same range for all three subgroups (**Figure 3)**. Post-boost IgG titers did not differ for previous SARS-CoV-2 infection, age, sex, and presence of any comorbidity (**Figure S1 and S2**).

**Figure 2.**
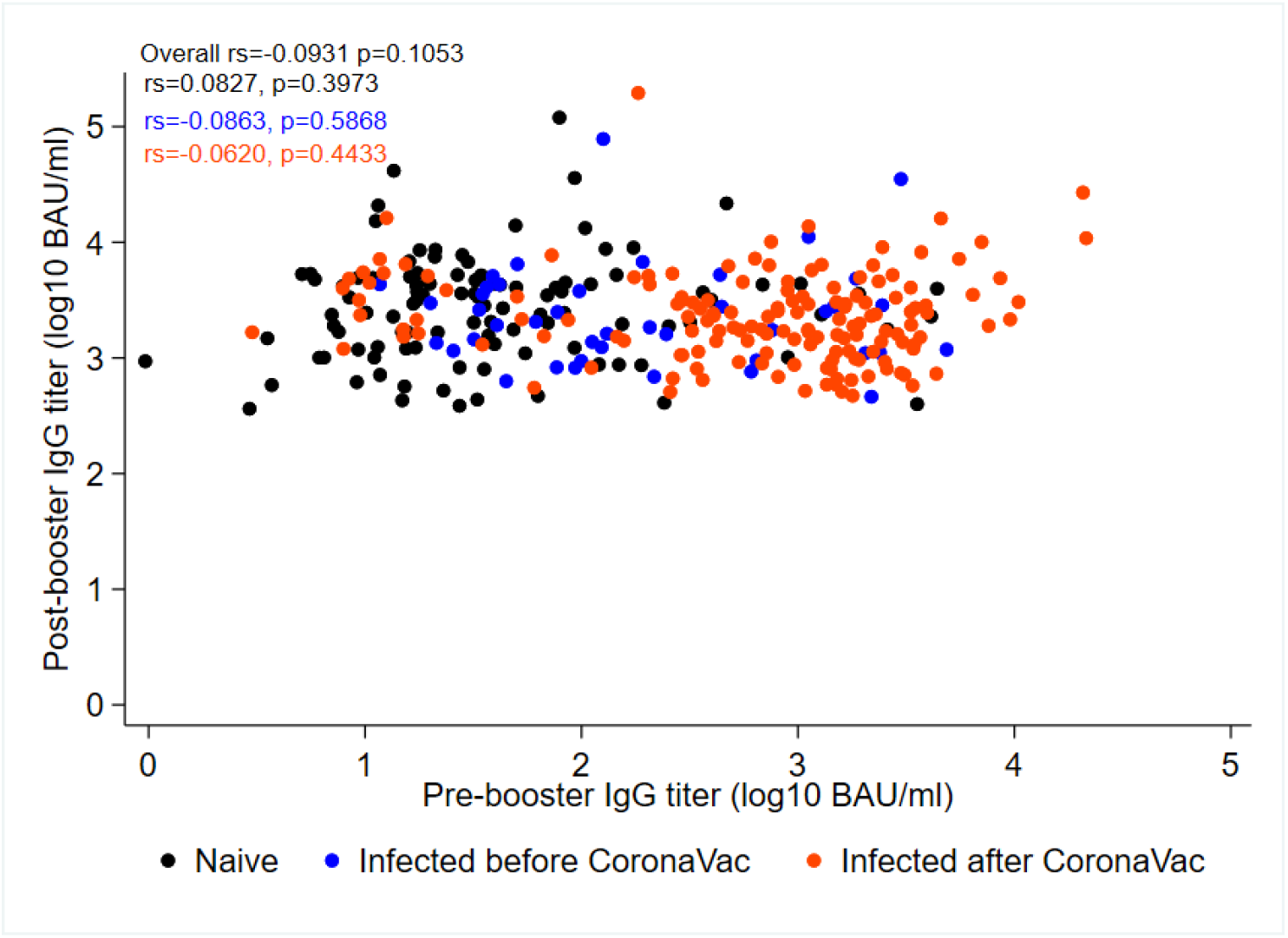
Scatter plot of anti-Spike IgG titers before and after mRNA-1273 booster. All participants reached a substantial post-boost titer, ranging between 2570 and 1,380,384 BAU/mL (2.73 log_10_ range), and this was irrespective of the pre-boost IgG titer, previous infection, or the timing of the previous infection (before or after two-dose CoronaVac vaccination). Participants with a breakthrough infection post-CoronaVac had a higher pre-boost IgG titer than those infected before Coronavac and the naïve participants. Post-boost IgG titers were in the same range for all 3 subgroups.

**Figure 3.**
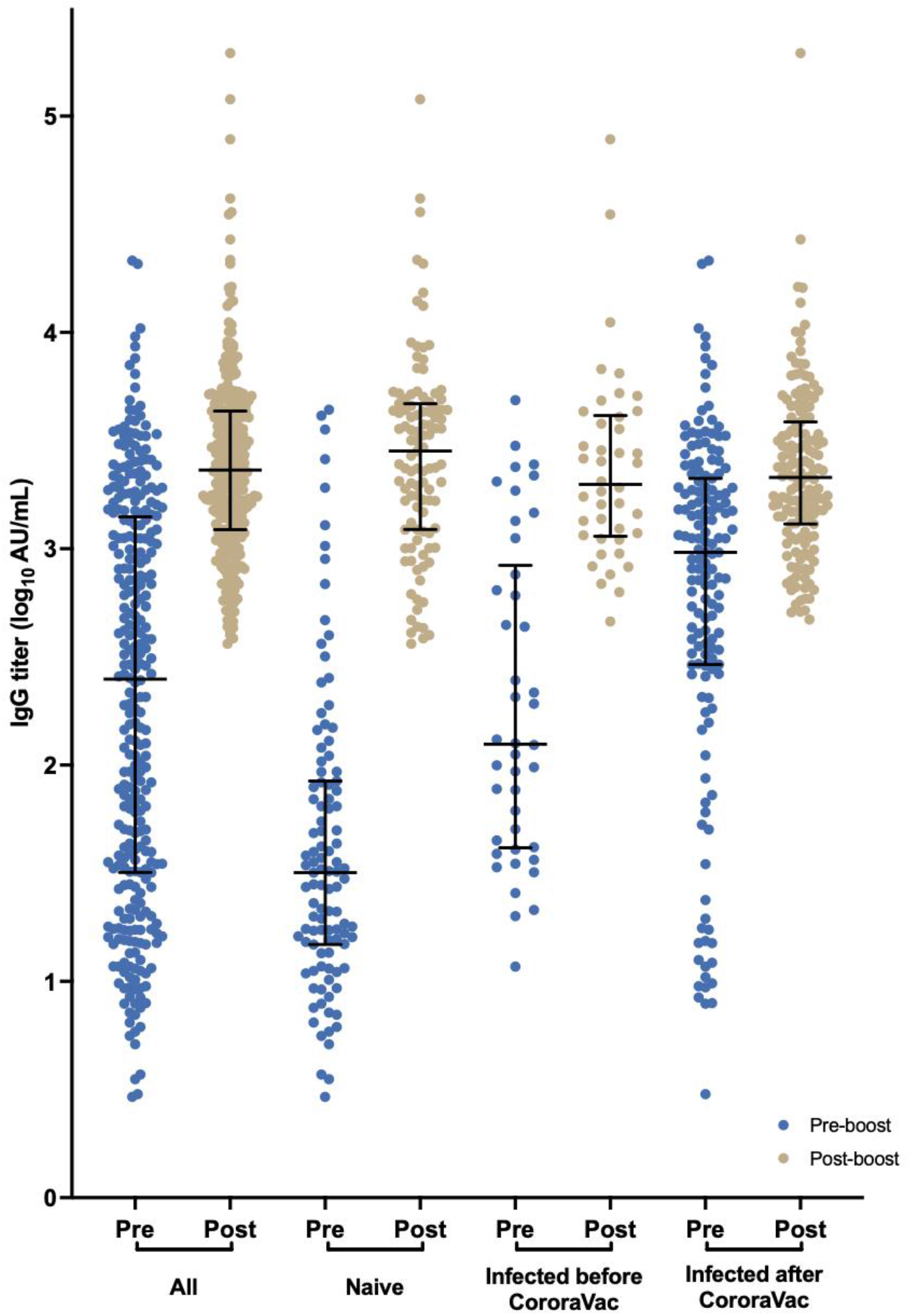
Anti-Spike IgG titers before and after mRNA-1273 booster. Dot plot showing pre and post-booster IgG titers, overall and by previous SARS-CoV-2 infection IgG titers shown as binding antibody units (BAU)/mL (according to WHO International Standard).

### Factors associated with pre- and post-booster IgG titers

In multivariable analysis, previous SARS-CoV-2 infection either before or after two-dose CoronaVac, and a longer time interval between second and mRNA-1273 boost dose were associated with a higher pre-boost IgG titer (p<0.0001 each); age, sex, comorbidity, time interval between first and second dose were not associated. None of these factors, including pre-boost IgG titer, were associated with a higher post-booster IgG titer (**Table 2**).

**Table 2.** Regression analysis of pre- and post-boost log_10_ anti-Spike IgG titers

|  | Pre-boost IgG |  |  |  |  |  | Post-boost IgG |  |  |  |  |  |
| --- | --- | --- | --- | --- | --- | --- | --- | --- | --- | --- | --- | --- |
|  | Univariable |  |  | Multivariable |  |  | Univariable |  |  | Multivariable |  |  |
|  | Beta | 95%CI | P-value | Beta | 95%CI | P | Beta | 95%CI | P-value | Beta | 95%CI | P |
| Age (per decade) | 0.095 | -0.094, 0.283 | 0.325 | 0.069 | -0.127, 0.264 | 0.491 | 0.111 | -0.079, 0.301 | 0.255 | 0.038 | -0.164, 0.239 | 0.713 |
| Sex, women | -0.230 | -0.717, 0.256 | 0.354 | -0.330 | -0.831, 0.171 | 0.306 | -0.015 | -0.500, 0.470 | 0.951 | -0.029 | -0.522, 0.463 | 0.907 |
| Comorbidity |  |  |  |  |  |  |  |  |  |  |  |  |
| Cardiovascular diseases | 0.418 | -0.475, 1.310 | 0.359 | - | - | - | 1.020 | 0.158, 1.882 | 0.020 | 0.816 | -0.110, 1.741 | 0.084 |
| Diabetes | -0.510 | -0.251, 1.489 | 0.617 | - | - | - | 2.360 | 0.133, 4.587 | 0.038 | 1.790 | -0.517, 4.097 | 0.128 |
| Obesity | 0.270 | -0.344, 0.885 | 0.388 | - | - | - | -0.163 | -0.763, 0.436 | 0.593 | - | - | - |
| Asthma | -0.990 | -2.316, 0.337 | 0.144 | - | - | - | 0.377 | -1.026, 1.780 | 0.599 | - | - | - |
| Previous SARS-CoV-2 infection |  |  |  |  |  |  |  |  |  |  |  |  |
| None | 1 (ref) |  |  |  |  |  | 1 (ref) |  |  |  |  |  |
| Before CoronaVac | **1.489** | **0.834, 2.144** | **<0.0001** | **1.938** | **1.232, 2.644** | **<0.0001** | -0.380 | -1.024, 0.264 | 0.248 | -0.288 | -0.952, 0.376 | 0.395 |
| After CoronaVac | **2.782** | **2.234, 3.331** | **<0.0001** | **2.404** | **1.824, 2.984** | **<0.0001** | -0.333 | -0.778, 0.113 | 0.143 | -0.155 | -0.693, 0.383 | 0.573 |
| Interval between first and second dose, day | -0.007 | -0.018, 0.006 | 0.296 | - | - | - | 0.001 | -0.013, 0.015 | 0.883 | - | - | - |
| Interval between second and mRNA-1273 booster dose, month | **0.453** | **0.318, 0.588** | **<0.0001** | **0.337** | **0.167, 0.506** | **<0.0001** | -0.018 | -0.147, 0.110 | 0.780 | - | - | - |
| Pre-boost IgG titer | - | - | - | - | - | - | -0.191 | -0.413, 0.032 | 0.094 | -0.163 | -0.432, 0.106 | 0.235 |

The figure shows results of univariable and multivariable ordinal regression analysis with pre-boost and post-boost IgG as the dependent variables (classified in quartiles).

### Reactogenicity of mRNA-1273 booster

The 304 participants reported a total of 300 adverse reactions within 7 days after mRNA-1273 booster. The percentage of participants with any solicited adverse reaction was 98.7% (300), 96.4% (293) for any local reaction (injection site pain and swelling most frequent) and 90.8% (276) for any systemic reaction (myalgia, fever/chills and headache being most frequent). Most solicited local and systemic adverse reactions were mild (86, 28.3%) or moderate (131, 43.1%), whereas severe (59, 19.4%) or those disrupting ADL (24, 7.9%) were less common (**Figure 4** and **Table S2**). All adverse reactions were short-lived and none required hospitalisation. Severe or ADL-disrupting adverse reactions were associated with longer interval between second dose and mRNA1273 booster (OR 1.37 per month increase, 1.09-1.72, p=0.001), and inversely associated with age (OR 0.71 per decade increase, 0.54-0.92, p=0.011); sex, comorbidity, previous SARS-CoV-2 infection, time interval between first and second dose, and pre- and post-boost IgG titer were not associated with any adverse reactions (**Table S3**).

**Figure 4.**
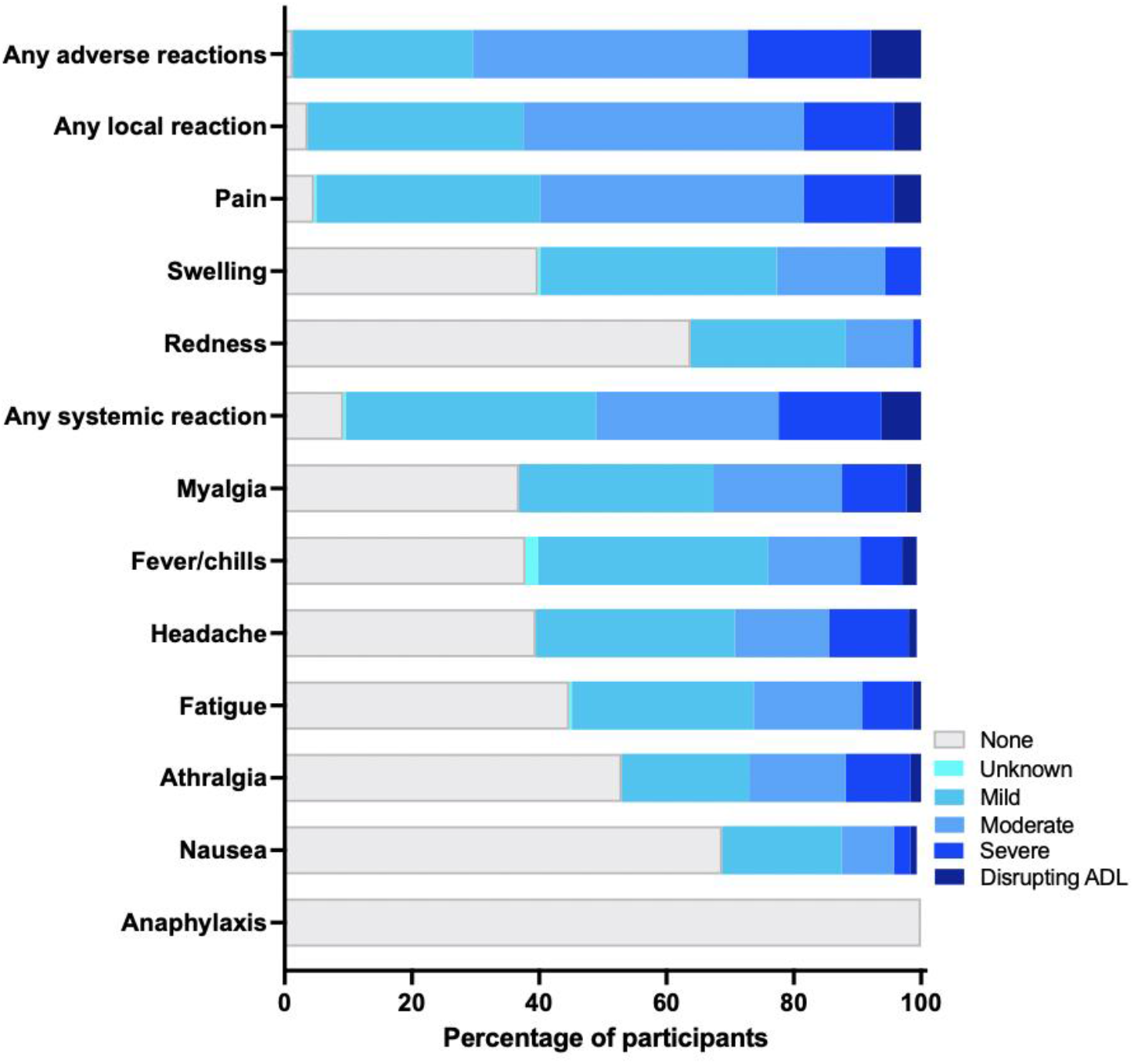
Adverse reactions within seven days after receiving the mRNA-1273 booster dose The bar chart shows solicited adverse reactions, that were verified and graded for severity by the study physician. Severity grading: 1) mild (does nor or minimally interfere with usual social and functional activities); 2) moderate (interferes with usual social and functional activities); 3) severe (causing inability to perform usual social and functional activities); 4) disrupting/impairing activities of daily living (ADL). No participants required hospitalization or died.

## Discussion

This real-world study found that a heterologous mRNA-1273 vaccine boost dose in Indonesian hospital staff with high SARS-CoV-2 exposure, after a primary course of two-dose CoronaVac six months prior, resulted in a median 9.3-fold increase in anti-spike serum IgG titers measured after 28 days, from a median of 250 (IQR 32-1389) to 2313 (IQR 1226-4324) BAU/mL. All participants, including seven individuals whose pre-boost IgG was below assay detection limits, became seropositive and all reached a substantial post-boost titer (≥364 BAU/mL). Post-boost IgG levels were not associated with pre-boost antibody titer or previous SARS-CoV-2 infection. Higher pre-boost IgG levels were associated with documented previous SARS-CoV-2 infection, and, unexpectedly, a longer interval since the second vaccine dose. However, when we assessed time since the most recent documented “virus exposure” (either by vaccine or infection), the pre-boost IgG titers correlated negatively, as expected^19–21^. These findings, coupled with the high rate of documented post-CoronaVac virus breakthroughs, confirm intense SARS-CoV-2 exposure among front line heath care workers during the Delta surge in July 2021^25^. Delta breakthrough infections have been associated with more potent, durable and resilient responses to spike mutations observed in variants of concern than those observed in subjects who were infected only or received only two doses of COVID-19 vaccine^28^.

The reactogenicity of the high-dose heterologous mRNA-1273 booster was acceptable, with common reports of injection site pain and swelling as well as muscle pain, fever/chills and headache, and 7.9% experienced a short-lived impairment in performing ADL. The booster was better tolerated with increasing age and a shorter time since the second dose. The findings largely concur with Moderna’s phase 2/3 trial of the high-dose (100μg) booster dose, which was generally safe and well-tolerated, although with slightly more frequent adverse reactions compared to the authorised 50μg dose^29^.

Emerging evidence for currently available vaccines suggests that heterologous boosting improves the breadth of humoral and cellular protection more than homologous boosting^11,12^. A small randomized trial in Hong Kong in adults who had received two-dose CoronaVac with low antibody response, found that an additional heterologous mRNA vaccine booster dose of BNT162b2 (Pfizer-BioNTech) was significantly more immunogenic than a homologous booster dose of CoronaVac booster against wild-type and VOCs^18^. Another study in heath care workers in Thailand who received two-dose CoronaVac found that a heterologous virus-vector booster AZD1222 (AstaZeneca) increased spike RBD-specific IgG 9-10-fold, with high neutralizing activity against the wild type and VOCs^26^.

Policy makers in several countries have started implementing third or periodic boosting to protect the most vulnerable populations, and mitigate health care and economic impacts. Real-world data and cohort studies are critical to guide decisions regarding when, which populations and what boosters should be administered. Compared to previous SARS-CoV-2 variants, current vaccine boosters appeared to neutralise Delta to a slightly lesser degree^12^, and Omicron to a substantially lesser degree^7^. Nonetheless, on 12^th^ December, Moderna announced preliminary data that the authorised booster dose of 50μg mRNA-1273 increased neutralising antibodies 37-fold and the high-dose of 100μg 83-fold, compared to pre-boost levels^29^. Further trials on clinical endpoints of optimized booster doses and variant-specific or multivalent vaccines in response to decreased susceptibility to neutralization of emerging SARS-CoV-2 VOCs are urgently needed^30^.

There are some limitations to our study. First, although accumulating evidence suggests that anti-Spike IgG response is a correlate of disease protection^31,32^, it is important to recognise there is not yet an established or well defined correlate of long-term vaccine protection. In this study we did not assess anti-N IgG, neutralizing antibody or cellular immunity^33^. Second, because subjects had not been under routine surveillance for SARS-CoV-2 infection, we could not fully discern all who had been infected by SARS-CoV-2, and the use of anti-N as a proxy for breakthrough infections was not valid because of the initial immunization with a whole-virus inactivated vaccine. Third, the sample size and follow-up period were not sufficient to identify less common or late adverse events following booster vaccination, and the immunogenicity data were limited to immune responses through study day 28. Fourth, only immunological data were collected and therefore this investigation lacks information regarding the efficacy of a heterologous mRNA-1273 booster vaccination. Lastly, demographics of the volunteers were not representative of the Indonesian population, and elderly were not represented.

In conclusion, our study provides evidence that, after a primary course of CoronaVac inactivated virus vaccine, a heterologous, high-dose mRNA-1273 booster was highly immunogenic and safe, even for those with the lowest pre-boost antibody levels.

## Data Availability

After publication, the datasets used for this study will be made available to others on reasonable requests to the corresponding author, including a detailed research proposal, study objectives and statistical analysis plan. Deidentified participant data will be provided after written approval from the principal investigators.

## Acknowledgements

The authors thank the study participants, the staff at the collaborating clinical sites and the Prodia laboratory.

## Authors’ contributions

EN and RLH are the INVITE principal investigators. RS, DU, EN, AS and RLH conceptialised the study. RS, DU, CN, F, DT, MR, AFR, established the cohort, collected the clinical samples and data. S and DS designed, supervised and performed the antibody testing. NN and WH managed the clinical database. HS performed the statistical analysis, with input from AS and RLH. RLH wrote the first draft of the manuscript. All authors provided valuable input to interpretation of the data and critically reviewed the paper and figures for important intellectual content. All authors reviewed and approved the final version of the manuscript.

## References

1 COVID-19 Vaccine Tracker. 2021. https://covid19.trackvaccines.org/agency/who/.

2 Levin EG, Lustig Y, Cohen C, et al. Waning Immune Humoral Response to BNT162b2 Covid-19 Vaccine over 6 Months. N Engl J Med 2021; published online Oct 6. DOI:10.1056/NEJMoa2114583.

3 Pouwels KB, Pritchard E, Matthews PC, et al. Effect of Delta variant on viral burden and vaccine effectiveness against new SARS-CoV-2 infections in the UK. Nat Med 2021. DOI:10.1038/s41591-021-01548-7.

4 Tartof SY, Slezak JM, Fischer H, et al. Effectiveness of mRNA BNT162b2 COVID-19 vaccine up to 6 months in a large integrated health system in the USA: a retrospective cohort study. Lancet (London, England) 2021; 398: 1407–16.

5 Chemaitelly H, Tang P, Hasan MR, et al. Waning of BNT162b2 Vaccine Protection against SARS-CoV-2 Infection in Qatar. N Engl J Med 2021; published online Oct 6. DOI:10.1056/NEJMoa2114114.

6 Lu L, Mok BW-Y, Chen L-L, et al. Neutralization of SARS-CoV-2 Omicron variant by sera from BNT162b2 or Coronavac vaccine recipients. Clin Infect Dis 2021; : ciab1041.

7 UK Health Security Agency. SARS-CoV-2 variants of concern and variants under investigation in England Technical briefing 31 10 December 2021. 2021.

8 Liu X, Shaw RH, Stuart AS V, et al. Safety and immunogenicity of heterologous versus homologous prime-boost schedules with an adenoviral vectored and mRNA COVID-19 vaccine (Com-COV): a single-blind, randomised, non-inferiority trial. Lancet (London, England) 2021; 398: 856–69.

9 Barros-Martins J, Hammerschmidt SI, Cossmann A, et al. Immune responses against SARS-CoV-2 variants after heterologous and homologous ChAdOx1 nCoV-19/BNT162b2 vaccination. Nat Med 2021; 27: 1525–9.

10 Nordström P, Ballin M, Nordström A. Effectiveness of heterologous ChAdOx1 nCoV-19 and mRNA prime-boost vaccination against symptomatic Covid-19 infection in Sweden: A nationwide cohort study. Lancet Reg Heal – Eur 2021; 11. DOI:10.1016/j.lanepe.2021.100249.

11 Atmar RL, Lyke KE, Deming ME, et al. Heterologous SARS-CoV-2 Booster Vaccinations -Preliminary Report. medRxiv Prepr Serv Heal Sci 2021; : 2021.10.10.21264827.

12 Munro APS, Janani L, Cornelius V, et al. Safety and immunogenicity of seven COVID-19 vaccines as a third dose (booster) following two doses of ChAdOx1 nCov-19 or BNT162b2 in the UK (COV-BOOST): a blinded, multicentre, randomised, controlled, phase 2 trial. Lancet 2021; published online Dec 6. DOI:10.1016/S0140-6736(21)02717-3.

13 Folegatti PM, Ewer KJ, Aley PK, et al. Safety and immunogenicity of the ChAdOx1 nCoV-19 vaccine against SARS-CoV-2: a preliminary report of a phase 1/2, single-blind, randomised controlled trial. Lancet 2020; 396: 467–78.

14 World Health Organization. WHO recommendation of Sinovac COVID-19 vaccine (Vero Cell [Inactivated]) – CoronaVac. 2021.

15 Tanriover MD, Doğanay HL, Akova M, et al. Efficacy and safety of an inactivated whole-virion SARS-CoV-2 vaccine (CoronaVac): interim results of a double-blind, randomised, placebo-controlled, phase 3 trial in Turkey. Lancet 2021; 398: 213–22.

16 Jara A, Undurraga EA, González C, et al. Effectiveness of an Inactivated SARS-CoV-2 Vaccine in Chile. N Engl J Med 2021; 385: 875–84.

17 Hitchings MDT, Ranzani OT, Scaramuzzini Torres MS, et al. Effectiveness of CoronaVac among healthcare workers in the setting of high SARS-CoV-2 Gamma variant transmission in Manaus, Brazil: A test-negative case-control study. medRxiv 2021; : 2021.04.07.21255081.

18 Mok CKP, Cohen CA, Cheng SMS, et al. Comparison of the immunogenicity of BNT162b2 and CoronaVac COVID-19 vaccines in Hong Kong. Respirology 2021; published online Nov. DOI:10.1111/resp.14191.

19 Sauré D, O’Ryan M, Torres JP, Zuniga M, Santelices E, Basso LJ. Dynamic IgG seropositivity after rollout of CoronaVac and BNT162b2 COVID-19 vaccines in Chile: a sentinel surveillance study. Lancet Infect Dis 2021; published online Dec 22. DOI:10.1016/S1473-3099(21)00479-5.

20 Lim WW, Mak L, Leung GM, Cowling BJ, Peiris M. Comparative immunogenicity of mRNA and inactivated vaccines against COVID-19. The Lancet Microbe 2021; 2: e423.

21 Zeng G, Wu Q, Pan H, et al. Immunogenicity and safety of a third dose of CoronaVac, and immune persistence of a two-dose schedule, in healthy adults: interim results from two single-centre, double-blind, randomised, placebo-controlled phase 2 clinical trials. Lancet Infect Dis 2021; published online Dec 19. DOI:10.1016/S1473-3099(21)00681-2.

22 Ranzani OT, Hitchings MDT, Dorion M, et al. Effectiveness of the CoronaVac vaccine in older adults during a gamma variant associated epidemic of covid-19 in Brazil: test negative case-control study. BMJ 2021; 374: 2015–n2015.

23 WHO. Meeting of the Strategic Advisory Group of Experts (SAGE) on Immunization 4-7 October 2021. 2021.

24 Indonesia: Coronavirus Pandemic Country Profile. Published online at OurWorldInData.org. 2021. https://ourworldindata.org/coronavirus/country/indonesia.

25 Surendra H, Salama N, Lestari KD, et al. Pandemic inequity in a megacity: a multilevel analysis of individual, community and health care vulnerability risks for COVID-19 mortality in Jakarta, Indonesia. medRxiv 2021; : 2021.11.24.21266809.

26 Yorsaeng R, Suntronwong N, Phowatthanasathian H, et al. Immunogenicity of a third dose viral-vectored COVID-19 vaccine after receiving two-dose inactivated vaccines in healthy adults. Vaccine 2021. DOI:https://doi.org/10.1016/j.vaccine.2021.11.083.

27 WHO. EXPERT COMMITTEE ON BIOLOGICAL STANDARDIZATION. Establishment of the WHO International Standard and Reference Panel for anti-SARS-CoV-2 antibody. 2021 https://cdn.who.int/media/docs/default-source/biologicals/ecbs/bs-2020-2403-sars-cov-2-ab-ik-17-nov-2020_4ef4fdae-e1ce-4ba7-b21a-d725c68b152b.pdf?sfvrsn=662b46ae_8&download=true.

28 Walls AC, Sprouse KR, Joshi A, et al. Delta breakthrough infections elicit potent, broad and durable neutralizing antibody responses. bioRxiv 2021; : 2021.12.08.471707.

29 Moderna announces preliminary booster data and updates strategy to address Omikron variant. News release on DSecember 12th, 2021. https://s29.q4cdn.com/745959723/files/doc_news/Moderna-Announces-Preliminary-Booster-Data-and-Updates-Strategy-to-Address-Omicron-Variant-2021.pdf.

30 Choi A, Koch M, Wu K, et al. Safety and immunogenicity of SARS-CoV-2 variant mRNA vaccine boosters in healthy adults: an interim analysis. Nat Med 2021; 27: 2025–31.

31 Harvey RA, Rassen JA, Kabelac CA, et al. Association of SARS-CoV-2 Seropositive Antibody Test With Risk of Future Infection. JAMA Intern Med 2021; 181: 672–9.

32 Feng S, Phillips DJ, White T, et al. Correlates of protection against symptomatic and asymptomatic SARS-CoV-2 infection. Nat Med 2021; 27: 2032–40.

33 Sahin U, Muik A, Derhovanessian E, et al. COVID-19 vaccine BNT162b1 elicits human antibody and TH1 T cell responses. Nature 2020; 586: 594–9.

