## Supplementary material for "Serum anti-Spike antibody titers before and after heterologous booster with mRNA-1273 SARS-CoV-2 vaccine following two doses of inactivated whole-virus CoronaVac vaccine": Suplementary figures and tables

**Figure S1. Scatter plot of anti-Spike IgG titers before mRNA-1273 booster and time since second vaccination**

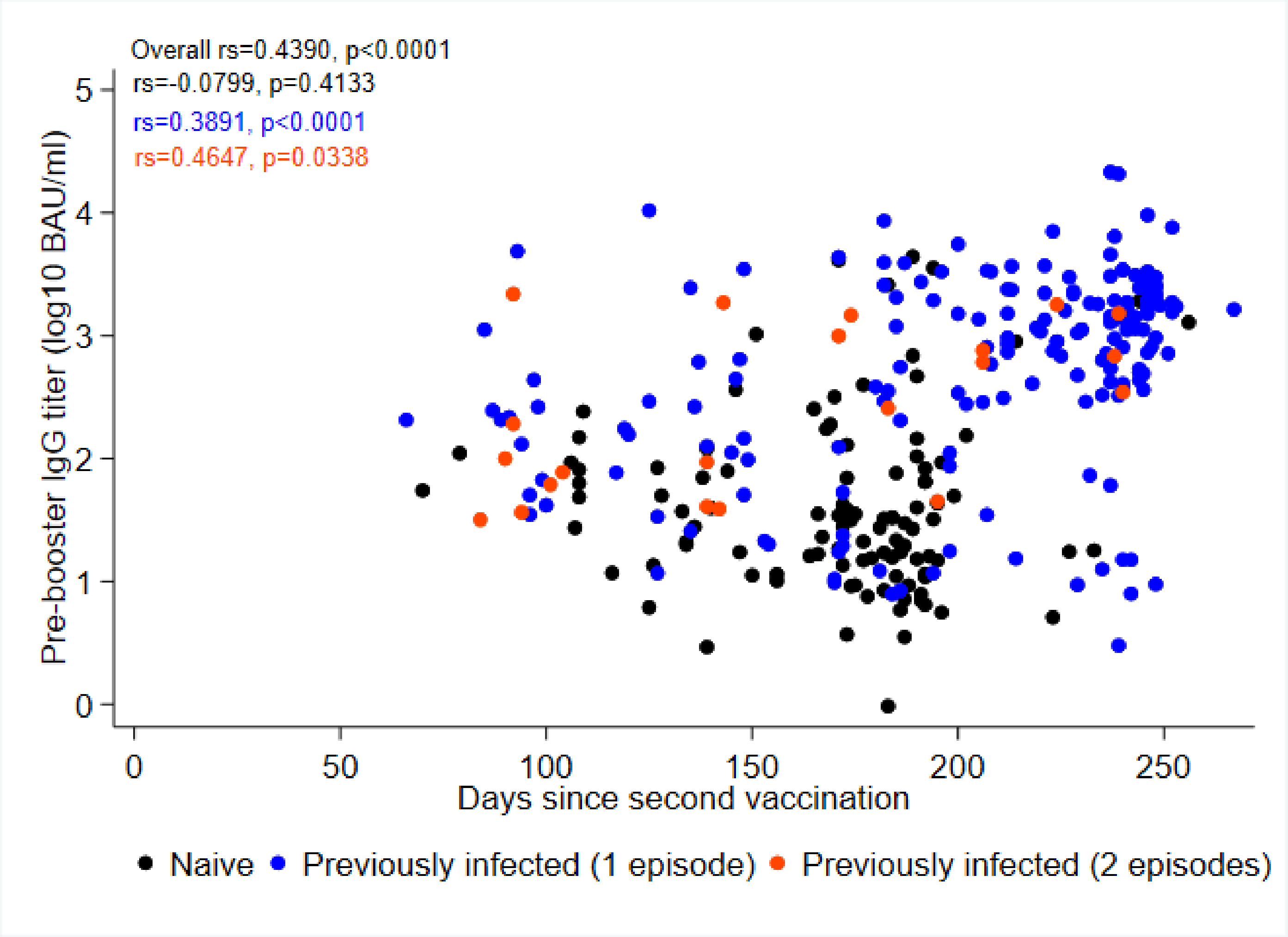


Pre-boost IgG titers correlated positively with the time since the second CoronaVac vaccine dose in previously infected participants (either one or two episodes), but did not correlate in naïve participants.

**Figure S2. Scatter plot of anti-Spike IgG titers before and after mRNA-1273 booster**

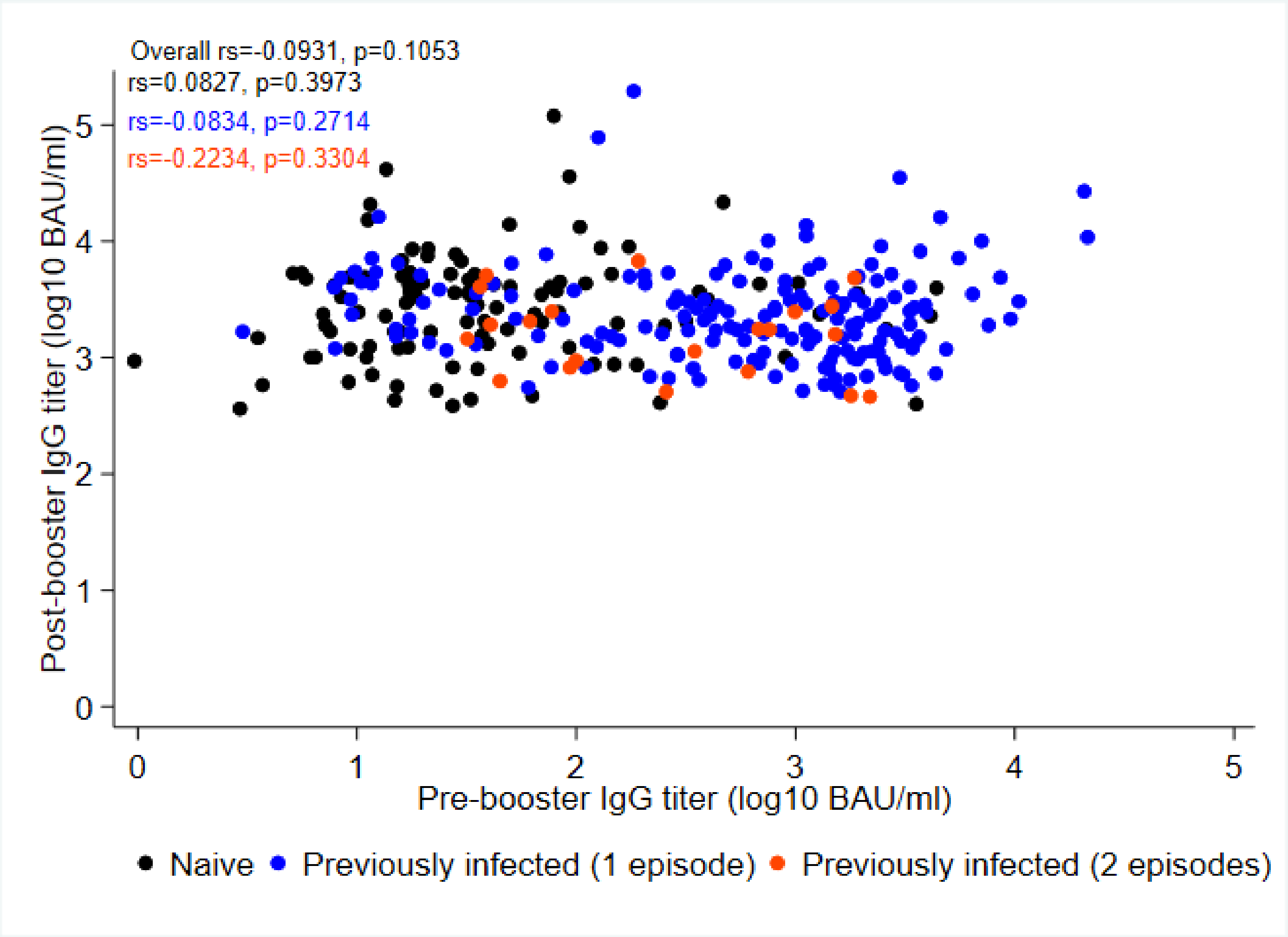


All participants reached a substantial post-boost titer, ranging between 2570 and 1,380,384 BAU/mL (2.73 $log_{10}$ range), and this was irrespective of the pre-boost IgG titer or previous infection.

**Figure S3. Anti-Spike IgG titers before and after mRNA-1273 booster**

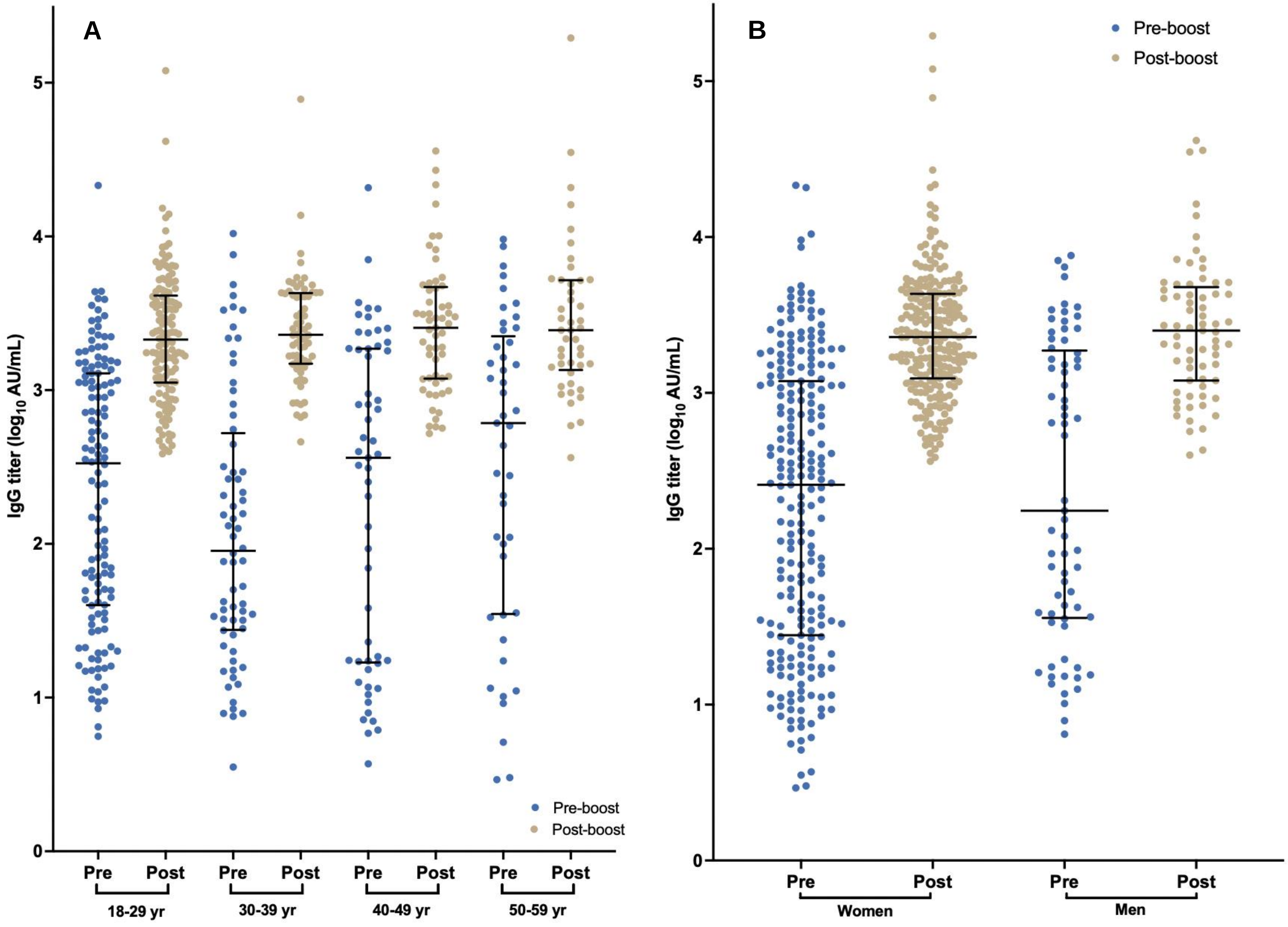

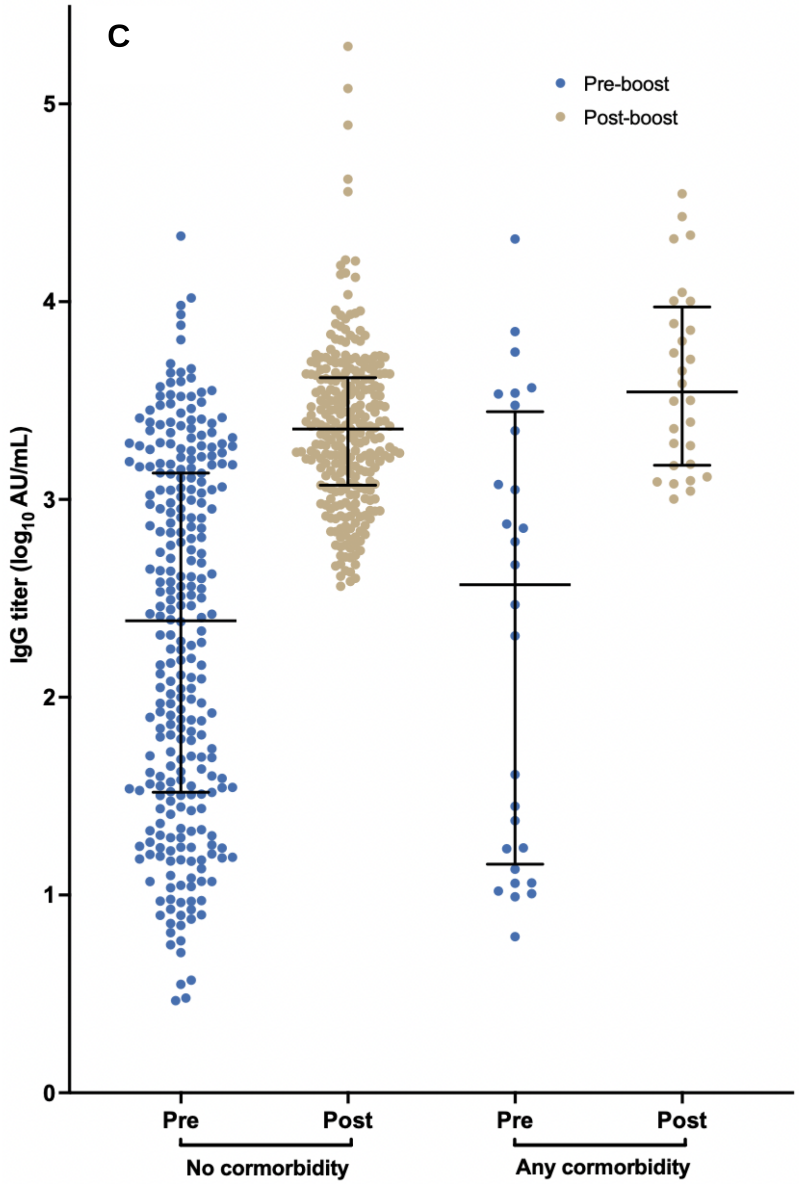


Panels (A-C) show dot plots of pre and post-booster IgG titers, by (A) age groups, (B) sex, (C) comorbidity.

IgG titers shown as binding antibody units (BAU)/mL (according to WHO International Standard).

**Figure S3. Change in anti-Spike IgG titers between before and after mRNA-1273 booster**

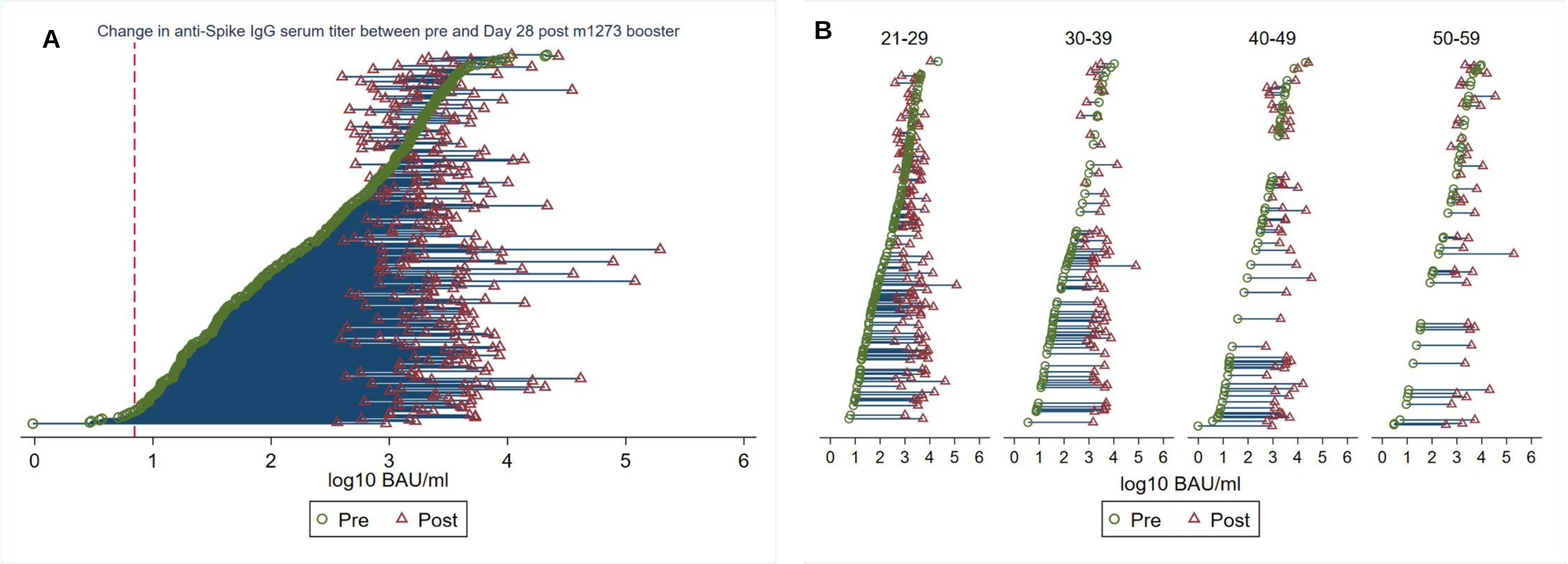

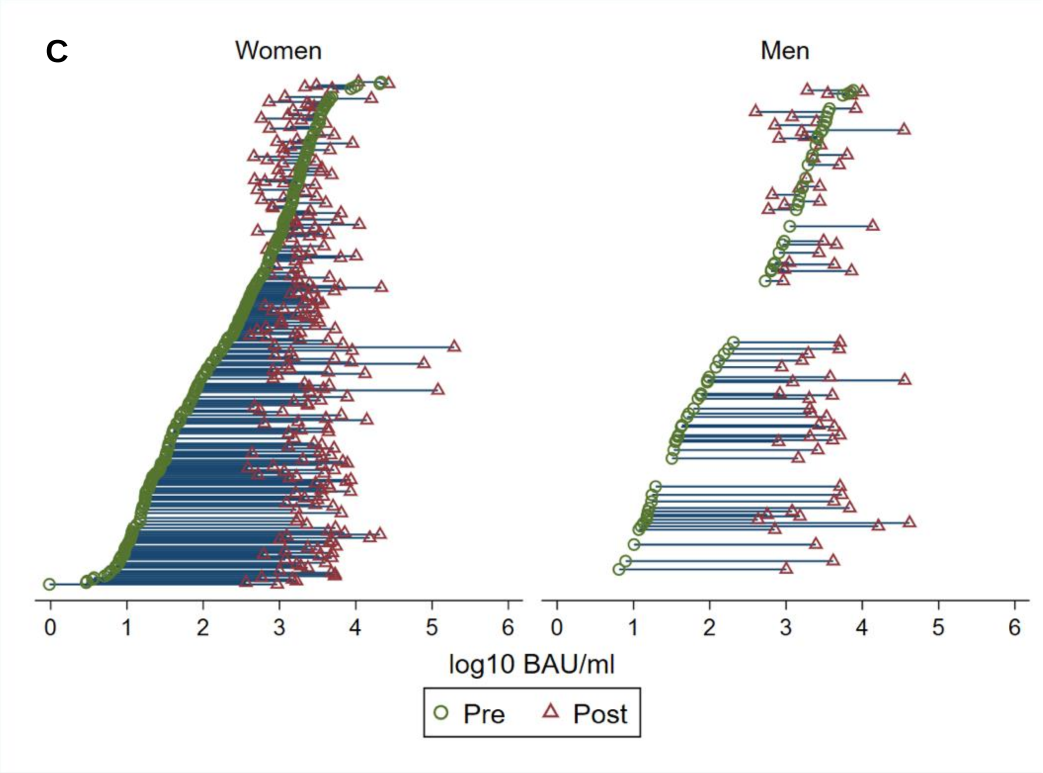


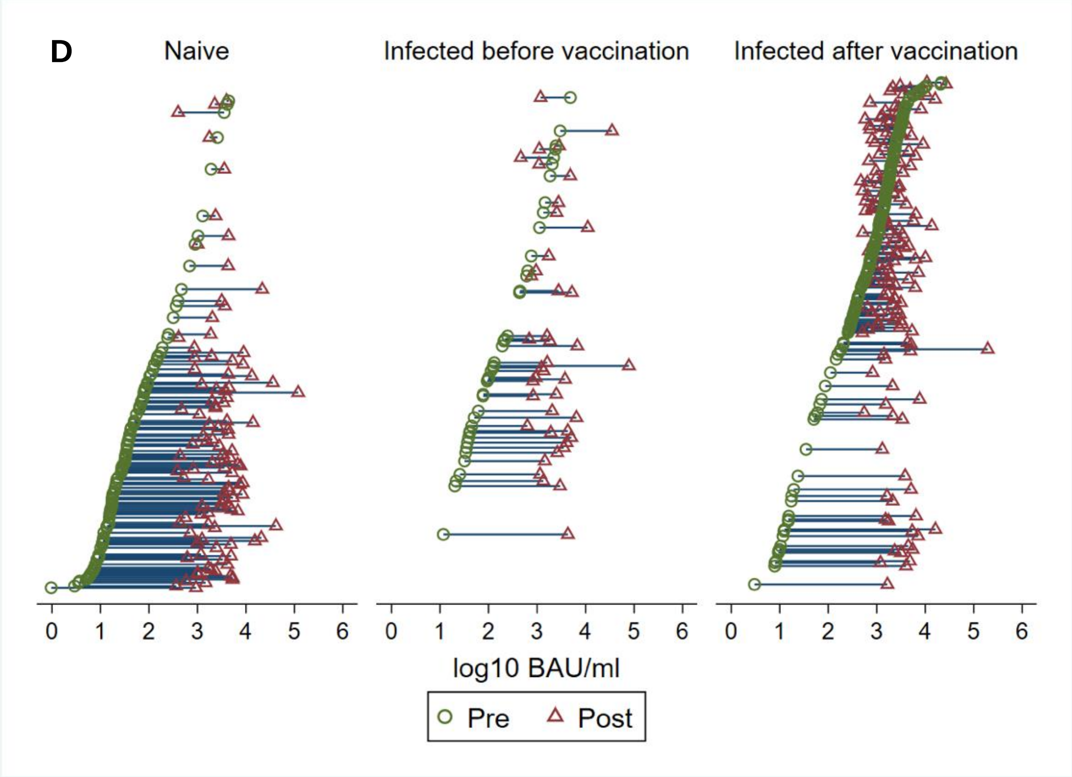


Panels (A-D) show the change in anti-Spike serum titer between pre and Day 28 post mRNA-1273 booster, (A) overall and by (B) age groups, (C) sex, (D) previous SARS-CoV-2 infection before or after CoronaVac. Participants (N=304) are ranked by pre-booster IgG titer**.** Dashed red line (A) represents cut-off of seropositivity (≥7.1 BAU/mL).

**Table S1. Anti-Spike IgG titers before and Day 28 after mRNA-1273 booster**

| | Median pre-booster (IQR) | Median post-booster (IQR) | Fold change | p-value |
|---|---|---|---|---|
| Overall | 249.8 (31.9-1389.2) | 2312.8 (1225.8-4324.3) | 9.3 | <0.0001 |
| Age | | | | |
| 21-29 | 334.9 (39.9-1285.1) | 2132.8 (1128.2-4066.2) | 6.4 | <0.0001 |
| 30-39 | 90.2 (27.7-500.4) | 2292.2 (1500.2-4271.4) | 25.0 | <0.0001 |
| 40-49 | 363.1 (17.1-1856.7) | 2543.9 (1197.1-4594.4) | 7.0 | <0.0001 |
| 50-59 | 611.0 (35.5-2048.7) | 2455.2 (1410.2-5177.3) | 2.8 | 0.0001 |
| Sex | | | | |
| Women | 257.1 (27.9-1191.1) | 2278.4 (1238.2-4315.4) | 8.9 | <0.0001 |
| Men | 175.2 (36.4-1804.3) | 2510.6 (1202.7-4570.3) | 14.3 | <0.0001 |
| Previous SARS-CoV2 infection | | | | |
| 0 | 31.8 (14.8-84.3) | 2833.6 (1227.6-4686.7) | 89.1 | <0.0001 |
| 1 | 808.5 (193.5-2049.1) | 2163.0 (1301.4-4033.3) | 2.7 | <0.0001 |
| 2 | 257.1 (61.5-994.1) | 1741.6 (823.6-2492.8) | 6.8 | 0.0010 |
| Previous SARS-CoV-2 infection timing | | | | |
| None | 31.8 (14.8-84.3) | 2833.6 (1227.6-4686.7) | 89.1 | <0.0001 |
| Before CoronaVac | 124.9 (41.7-760.3) | 1985.5 (1153.0-4077.5) | 15.9 | <0.0001 |
| After CoronaVac | 962.6 (292.0-2115.5) | 2136.4 (1299.3-3854.6) | 2.2 | <0.0001 |
| Time interval between second dose and mRNA-1273 booster | | | | |
| 1 | 80.1 (34.3-211.6) | 2369.6 (1202.0-4324.3) | 24.6 | <0.0001 |
| 2 | 33.0 (15.5-294.0) | 2943.0 (1482.5-4686.7) | 94.6 | <0.0001 |
| 3 | 709.4 (49.6-1596.1) | 2522.3 (1303.6-5305.1) | 3.4 | <0.0001 |
| 4 | 1439.5 (533.8-2226.6) | 1714.7 (1132.5-2764.0) | 1.2 | 0.0037 |
| Comorbidity | | | | |
| 0 | 243.9 (33.1-1359.9) | 2269.9 (1180.7-4115.5) | 9.3 | <0.0001 |
| ≥1 | 380.9 (15.3-2608.7) | 3510.3 (1494.9-8898.8) | 9.2 | <0.0001 |

Data are expressed as binding antibody units (BAU)/mL (according to WHO International Standard)

**Table S2**. Adverse reactions within seven days after receiving the mRNA-1273 booster dose

| | **N (%)** | **Grade 1** | **Grade 2** | **Grade 3** | **Grade 4** | **Unknown** |
|---|---|---|---|---|---|---|
| Any adverse reactions | 300 (98.68) | 86 (28.29) | 131 (43.09) | 59 (19.41) | 24 (7.89) | 0 (0.00) |
| Any local reaction | 293 (96.38) | 103 (33.88) | 134 (44.08) | 43 (14.14) | 13 (4.28) | 0 (0.00) |
| Pain | 290 (95.39) | 107 (35.20) | 126 (41.45) | 43 (14.14) | 13 (4.28) | 1 (0.33 |
| Swelling | 183 (60.20) | 113 (37.17) | 52 (17.11) | 17 (5.59) | 0 (0.00) | 1 (0.33) |
| Redness | 110 (36.18) | 74 (24.34) | 32 (10.53) | 4 (1.32) | 0 (0.00) | 0 (0.00) |
| Any systemic reaction | 276 (90.79) | 120 (39.47) | 87 (28.62) | 49 (16.12) | 19 (6.25) | 1 (0.33) |
| Myalgia | 192 (63.16) | 93 (30.59) | 61 (20.07) | 31 (10.20) | 7 (2.30) | 0 (0.00) |
| Fever/chills | 189 (62.17) | 110 (36.18) | 44 (14.47) | 20 (6.58) | 7 (2.30) | 6 (1.97) |
| Headache | 184 (60.53) | 95 (31.25) | 45 (14.80) | 38 (12.50) | 4 (1.32) | 0 (0.00) |
| Fatigue | 168 (55.26) | 87 (28.62) | 52 (17.11) | 24 (7.89) | 4 (1.32) | 1 (0.33) |
| Arthralgia | 143 (47.04) | 61 (20.07) | 46 (15.13) | 31 (10.20) | 5 (1.64) | 0 (0.00) |
| Nausea | 95 (31.25) | 57 (18.75) | 25 (8.22) | 8 (2.63) | 3 (0.99) | 0 (0.00) |
| Anaphylaxis | 0 (0.0) | 0 (0.00) | 0 (0.00) | 0 (0.00) | 0 (0.00) | 0 (0.00) |

The table shows solicited adverse reactions, that were verified and graded for severity by the study physician. Severity grading: 1) mild (does nor or minimally interfere with usual social and functional activities); 2) moderate (interferes with usual social and functional activities); 3) severe (causing inability to perform usual social and functional activities); 4) disrupting/impairing activities of daily living (ADL). No participants required hospitalization or died.

**Table S3. Regression analysis of occurrence of severe or disrupting adverse reactions.**

| | Univariable | | | Multivariable | | |
|---|---|---|---|---|---|---|
| | **OR** | **95%CI** | **P** | **OR** | **95%CI** | **P** |
| Age, per decade increase | 0.69 | 0.53-0.90 | 0.006 | **0.71** | **0.54-0.92** | **0.011** |
| Sex | | | | | | |
| Men | 1 (ref) | | | 1 (ref) | | |
| Women | 1.63 | 0.85-3.130 | 0.139 | 1.60 | 0.81-3.17 | 0.174 |
| Comorbidities | | | | | | |
| No | 1 (ref) | | | - | - | - |
| Yes | 0.88 | 0.36-2.15 | 0.774 | - | - | - |
| Past COVID-19 | | | | | | |
| No | 1 (ref) | | | 1 (ref) | | |
| Yes | 1.73 | 0.99-3.03 | 0.053 | 1.20 | 0.59-2.44 | 0.619 |
| Pre-booster IgG | 1.43 | 1.08-1.89 | 0.013 | 0.73 | 0.33-1.61 | 0.431 |
| Post-booster IgG | 0.58 | 0.31-1.10 | 0.095 | - | - | - |
| Change in IgG titers between before and after mRNA-1273 boost | 0.69 | 0.53-0.89 | 0.004 | 0.65 | 0.33-1.30 | 0.225 |
| Interval between first and second vaccination, per day increase | 0.97 | 0.93-1.00 | 0.059 | 0.99 | 0.96-1.02 | 0.484 |
| Interval between second vaccination and mRNA-1273 booster dose, per month increase | **1.51** | **1.25-1.83** | **<0.0001** | **1.37** | **1.09-1.72** | **0.007** |

Table shows univariable and multivariable logistic regression with occurrence of any severe or ADL-disrupting adverse reaction as the dependent variable.

Severe or disrupting adverse reactions were defined as causing inability to perform usual social and functional activities or disrupting/impairing activities of daily living (ADL), respectively.

Interval between first and second dose, post-booster IgG titer, and presence of any comorbidity were not associated with the occurrence of severe or disrupting adverse reaction.
